# Implementation strategies for telemental health: a systematic review

**DOI:** 10.1101/2022.04.29.22274367

**Authors:** Rebecca Appleton, Phoebe Barnett, Norha Vera San Juan, Elizabeth Tuudah, Natasha Lyons, Jennie Parker, Emily Roxburgh, Spyros Spyridonidis, Camilla Tamworth, Minnie Worden, Melisa Yilmaz, Nick Sevdalis, Brynmor Lloyd-Evans, Justin J Needle, Sonia Johnson

## Abstract

**Background:** The COVID-19 pandemic resulted in a rapid shift from traditional face-to-face care provision towards delivering mental health care remotely through telecommunications, often referred to as telemental health care. However, the manner and extent of telemental health implementation have varied considerably across settings and areas, and substantial barriers are encountered. There is, therefore, now a need to identify what works best for service users and staff and establish the key mechanisms for efficient integration into routine care.

**Objective:** We aimed to identify investigations of pre-planned strategies intended to achieve or improve effective and sustained implementation of telemental health approaches, and to evaluate how different strategies influence implementation outcomes.

**Methods:** A systematic review was conducted, with five databases searched for relevant literature using any methodological approach, published between January 2010 and July 2021. Studies were eligible for inclusion if they took place in secondary or tertiary mental health services and focused on pre-planned strategies for achieving or improving delivery of mental health care through remote communication between mental health professionals or between mental health professionals and service users, family members, unpaid carers, or peer supporters. All included studies were assessed for risk of bias. Data were synthesised using the Expert Recommendations for Implementing Change (ERIC) compilation of implementation strategies and the taxonomy of implementation outcomes.

**Results:** A total of 14 studies were identified which met the inclusion criteria. A variety of implementation strategies were identified, the most commonly reported being ‘Train and educate stakeholders’. All studies reported using a combination of several implementation strategies.

**Conclusions:** Using a combination of implementation strategies appears to be a helpful method of supporting the implementation of telemental health. Further research is needed to test the impact of specific implementation strategies on implementation outcomes.

## Introduction

Telemental health (TMH) refers to delivering mental health care remotely via telecommunications technology (as opposed to face-to-face) (1). Adoption of TMH has expanded during the COVID-19 pandemic to allow services to continue to offer mental health support despite social distancing restrictions. This shift in care delivery was often conducted rapidly as part of the emergency response to the pandemic, in which services had to adapt their existing face-to-face models of treatment to include remote forms of care (2).

Some benefits to delivering mental health support in this way have been identified, for example, increasing access for service users who live remotely, have difficulty travelling, or find mental health care settings stigmatising or intimidating, and greater convenience for some service users (3, 4).

However, there are also some challenges associated with this approach, for example, some service users may not have access to technology, internet connectivity, or a private space to use during TMH care, whilst others have identified challenges in developing and maintaining a therapeutic relationship (5, 6). A recent systematic review also identified that TMH may not be suitable for all types of therapy, for example, exposure therapy or when treating trauma (2). This review also identified challenges in delivering TMH care to certain populations, for example, children and service users with learning difficulties or severe anxiety (2).

The rapid switch to TMH during the pandemic has resulted in great variations in how and to what extent TMH has been adopted and sustained, across different geographical locations and services (7). Due to the rapid nature of the implementation of TMH, staff have raised concerns around a lack of appropriate training to be able to conduct remote mental health care effectively and safely (6, 8). Nonetheless, both staff and service users express interest in incorporating TMH in routine care beyond the pandemic, increasing service user and staff choice and convenience. To move beyond the piecemeal pandemic implementation of TMH to strategies for incorporating it in routine care in the future, we need a greater understanding of the best approaches to introducing and sustaining it in contexts where it is potentially helpful.

Implementation strategies represent “the ‘how to’ component of changing healthcare practice” (9) and are key in determining the success of an intervention. There is a need to establish what works for whom in TMH and identify the key mechanisms for acceptable, effective and efficient integration into routine care. Studies using implementation science methods are especially focused on meeting this need. In our previous umbrella review of the literature predating the pandemic (10), we did not find any systematic reviews which focused on implementation strategies. A recent review conducted by James et al (11) which explored the implementation of video consultations in healthcare in general found that helpful facilitating strategies included the use of a telehealth champion and embedded leadership, whereas challenges included lack of technical experience among staff, a lack of perceived demand amongst staff or patients or motivation to change, a lack of a strategic plan for implementation, and cost issues. Whilst some studies of mental health services were included in this review, there is a need for a specific evidence base for mental health care, given the particular relational and risk challenges in this setting.

The current review therefore aimed to synthesise evidence on how best to implement and sustain TMH during the recovery from the pandemic and beyond, integrating it across the mental health system in a flexible and sustainable way that both maximises its potential in everyday practice and allows a response to be rapidly mobilised to any future emergency.

This review aims to:

1. identify and describe strategies that have been used to improve the implementation of TMH approaches;
2. synthesise evidence on how these strategies influence implementation outcomes.

We hope that the findings from this research will inform future service development as services adapt to the ‘new normal’ way of working following the COVID-19 pandemic, and support the identification of research questions and approaches for future investigations in this area.

## Methods

This systematic review is reported according to the Preferred Reporting Items for Systematic Reviews and Meta-Analyses (PRISMA) guidance (12). The review was prospectively registered on PROSPERO (CRD42021266245).

### Inclusion criteria

We included studies meeting the following criteria:

#### Participants

Staff who worked within secondary or tertiary mental health care settings (community and inpatient care); people of any age who received organised mental health care in secondary or tertiary mental health care settings, or their family members or carers.

Studies conducted in primary care or standalone psychotherapy service settings, or that involved service users with substance misuse, neuropsychiatry/neurology or dementia diagnoses were excluded.

#### Interventions

Pre-planned strategies to support the effective and sustained implementation of TMH within secondary or tertiary mental health care settings. We included all modalities of TMH, including video calls, telephone calls, text messaging platforms and hybrid approaches combining different platforms, or remote with face-to-face care. TMH care must have included spoken or written communication carried out remotely between mental health professionals or between mental health staff and patients, service users, family members, unpaid carers, or peer supporters.

Studies where the intervention was only delivered to selected participants recruited for the purpose of the study, as opposed to being rolled out across an existing service, were excluded.

#### Outcomes

At least one of the outcomes from Proctor and colleagues’ (13) taxonomy of implementation outcomes, defined for the purposes of this review as the effects of deliberate and purposive actions to implement TMH (see Evidence Synthesis for more detail), had to be reported:

- Acceptability (to service users or staff)
- Adoption (including any individual differences in those reached or not reached)
- Appropriateness
- Feasibility, e.g. actual fit, suitability for use
- Fidelity
- Cost and cost effectiveness (of implementation support intervention or strategy)
- Penetration, e.g. spread, level of institutionalisation
- Sustainability

Studies with or without a comparator were included. Studies were excluded which reported findings about the extent of implementation of a TMH programme, or described barriers to or facilitators of the implementation of TMH, but did not describe and evaluate an explicit pre-planned strategy designed to achieve more widespread, effective and/or sustained implementation of TMH, or did not report a relevant outcome according to Proctor’s taxonomy.

#### Study designs

There were no restrictions based on study design or language of papers.

### Further exclusion criteria

We also excluded conference abstracts, review articles, editorials and opinion pieces. Papers were excluded if they were published before January 2010 as earlier studies may be less relevant due to changes in both the availability of and familiarity with technology that can be used to support telehealth.

### Search strategy

The search strategy included the following:

1. Five academic databases (PubMed, EMBASE, PsycINFO, CINAHL and Web of Science) were searched from January 2010 to July 2021. The search strategy used a combination of keyword and subject heading searches relating to mental illness, remote working and implementation.
2. Preprint servers (medRxiv, PsyArXiv, Wellcome Open Research and JMIR Preprints) were searched (October 2021).
3. Forward citation searching using Web of Science and backward citation searching of reference lists of included studies.

The full search strategy is provided in Appendix 1.

### Screening

All references were de-duplicated in Endnote X9 (14) and then imported into Rayyan (15) for title and abstract screening. Title and abstract screening was conducted by five reviewers (RA, PB, SS, ER, CT), with 100% included references and 25% of excluded references checked by another member of the research team (PB, MY, MW) to ensure inclusion criteria had been applied correctly. Full text screening was conducted by four reviewers (PB, RA, ET, NL), with 100% included references and 25% of excluded references checked by another member of the research team. All disagreements were resolved by discussion with a third reviewer.

### Data extraction

All included references were imported into EPPI-Reviewer 4.0 (16) for data extraction. A data extraction form was created and piloted on a small number of included studies by three reviewers (RA, NSJ, PB), before data for the remaining studies was extracted by three reviewers (NL, ER, ET). All data extraction was checked by another member of the research team (PB, NSJ, RA, JP).

Details on the service setting, study design, characteristics of the clinical population, characteristics of the staff and TMH modalities used were extracted from each study. Details of the implementation strategy used, categorised according to the ERIC compilation of implementation strategies (17) (outlined in more detail below) and implementation outcomes (categorised according to Proctor’s taxonomy (13)) were also extracted.

### Quality appraisal

Quality appraisal was conducted using the Mixed Methods Appraisal Tool (MMAT) (18) for all primary research studies which aimed to answer a research question, and using the AACODS (Authority, accuracy, coverage, objectivity, date, significance) tool for descriptive studies (19) Quality assessment was carried out by one member of the research team (NL, ER, ET, or RA) and checked by another reviewer (RA, PB, or NVSJ). Due to the relatively small number of papers included in this review, the results of quality assessment were not used to determine eligibility for inclusion, although they were taken into account during interpretation of findings.

### Evidence synthesis

We conducted a framework synthesis (20) to consolidate findings from the included studies. The framework used for the synthesis was developed from two established implementation science frameworks. Firstly, we used the overarching categories from the ERIC compilation of implementation strategies (17), to identify and record strategies used to implement TMH (see Box 1 for further details). Secondly, outcomes of studies were categorised according to the Proctor taxonomy of implementation outcomes (13), which consists of the following: acceptability (stakeholders’ perception that the intervention is agreeable), adoption (uptake of the intervention), appropriateness (the perceived compatibility of the intervention), feasibility (the extent a new intervention can be used in a particular setting), fidelity (whether an intervention was implemented as originally prescribed), implementation cost (the cost effect of implementing the intervention), penetration (the integration of the intervention within a service) and sustainability (the extent to which a new intervention is maintained). Details of outcomes and interventions from the framework were used to create summaries of the strategies employed and resulting outcomes in each study. As per recommendations for the use of these outcomes (13), we recorded within the taxonomy both those outcomes reported as resulting from the pre-planned strategies intended to optimise TMH, and outcomes relating to the TMH interventions themselves (as a result of implementation strategies) where reported by the included studies. For example, we recorded acceptability of training reported by clinicians as well as acceptability of the TMH interventions reported by service users, which may have been impacted by the implementation strategy of train clinicians.

#### Box 1: Details of all categories covered by the ERIC compilation of implementation strategies *Use of evaluative and iterative strategies*

Examples include conducting a local needs assessment, provision of consumer feedback on the implementation, assessment for readiness, identification of barriers, quality monitoring tools, audit and feedback.

**Provision of interactive assistance**

Examples include processes of enabling and supporting individuals, groups, or organisations to adopt or incorporate effective practice, local technical assistance, ongoing supervision, and centralisation of technical assistance for implementation issues such as help-desks and online “frequently asked questions”.

**Adaptation and tailoring to context**

Examples include adapting interventions to address previously identified barriers, and identifying which aspects of the intervention can be adapted to suit need.

**Development of stakeholder interrelations**

Examples include identification of champions or leaders to support and drive implementation and overcome resistance, development of multi-disciplinary support teams with protected time to reflect on practice and share lessons, recruitment and cultivation of relationships with partners or community resources, such as charities, and identification of early adopters who others can learn from.

**Training and educating stakeholders**

Examples include ongoing training throughout implementation for clinicians, support staff and facilitators, ongoing consultation with experts, development of manuals and toolkits and training designated people to train others.

**Supporting clinicians**

Examples include facilitating the relay of information to clinicians, resource sharing agreements with organisations that have relevant required resources, revision of professional roles and changes to clinical teams to ensure the necessary skills are available.

**Engagement of consumers**

Examples include involving service users in the implementation effort, encouraging adherence, problem solving and spreading the word about the intervention.

**Utilising financial strategies**

Examples include funding to encourage uptake or incentivising adoption.

**Changes to infrastructure**

Examples include encouraging leadership to declare the intervention a priority, adaptation of physical structures such as room layout and changing accreditation and certification requirements.

## Results

### Study selection

Database searches identified 20,858 papers, of which 14,294 were screened by their title and abstract once duplicates had been removed. A total of 338 papers were screened at full text, resulting in 14 studies identified for inclusion in the review. No additional papers were identified from preprint servers or from forward or backward citation searching. The study selection and screening process is summarised in Figure 1.

**Figure 1:**
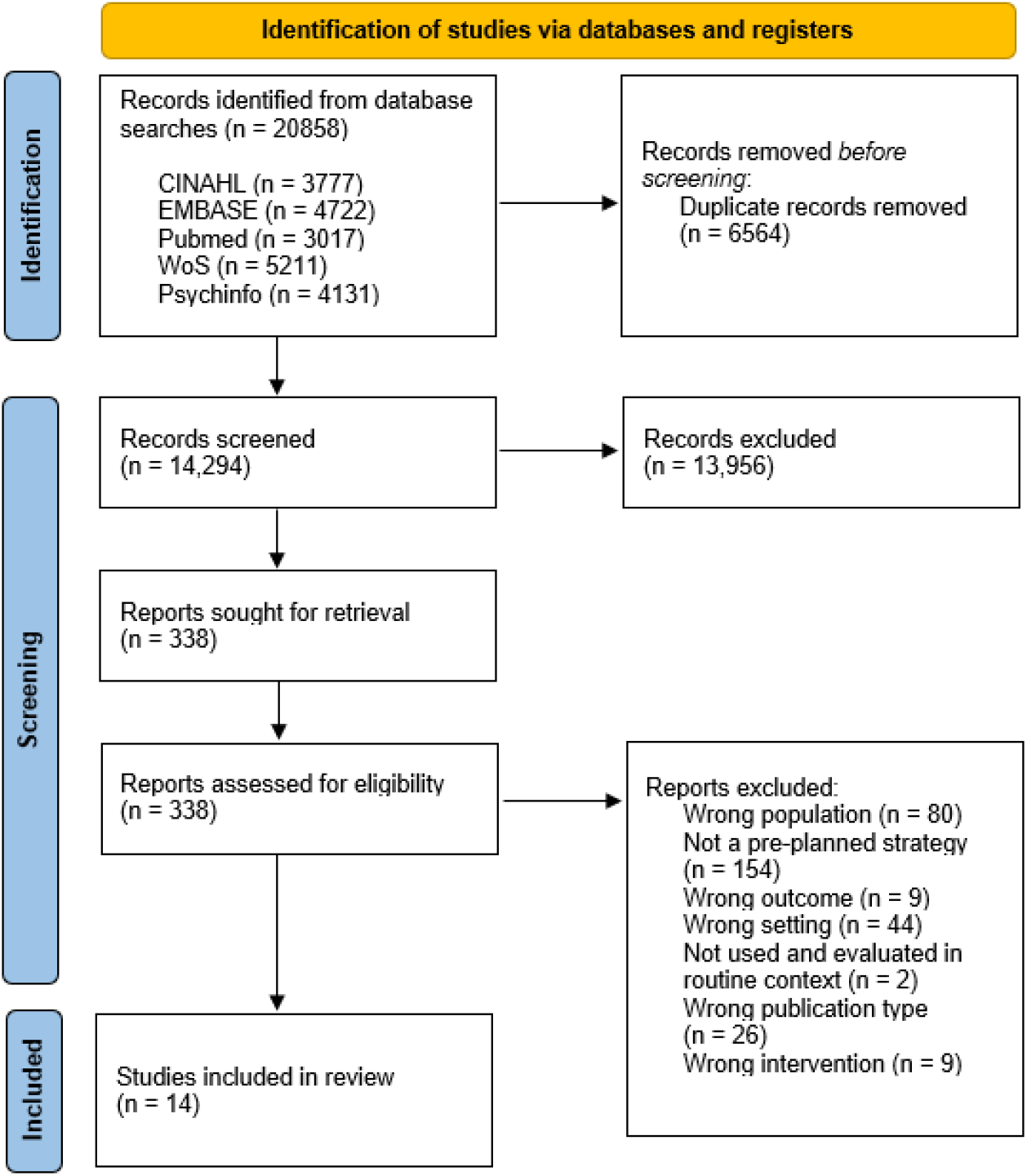
PRIMSA flow chart showing the study selection and screening process

### Study characteristics

Of the 14 included studies, 11 were conducted in the USA, with one each in the UK, Canada, and Australia. Six studies focused only on service users, seven only on staff, and one involved both staff and service users as participants. Four studies used quantitative methods, four were mixed methods studies, two were qualitative studies and four studies were descriptive in nature. No trials or studies with a comparison group were identified.

The majority (n=11) of studies explored implementation in community mental health services, and five studies used an established implementation framework to inform their analysis. Further details of included studies can be found in Table 1.

**Table 1:** Characteristics of included studies

| Author (year) | Country | Service Type | Aims | Study design | Participants: staff/ service-users, demographics n (%) or mean (SD) | TMH modalities | Theoretically informed implementation Framework used |
| --- | --- | --- | --- | --- | --- | --- | --- |
| Adler et al (21) | USA | Community mental health teams (CMHTs) and outpatient services, Veterans Affairs (VA) service | To examine changes in attitudes and knowledge regarding TMH following a pilot TMH service delivery improvement project and identify barriers and facilitators to its implementation in a VA service. | Pre-post pilot training programme | **Staff** (N=12)<br>*Job title n (%)*: Psychologists 7 (58), social workers 3 (25), other backgrounds 2 (17)<br>*Gender n (%)*: Male 4 (33), Female 8 (67);<br>Age: mean 44.6 | Video call | None stated |
| Baker-Ericzén et al (22) | USA | CMHTs and outpatient services | To describe the feasibility and acceptability of using a culturally adapted telemedicine intervention (the Perinatal Mental Health model) to ameliorate the barriers to adequate diagnosis and intervention for maternal depression in Latina women | Feasibility pilot study | **Service users** (N=79)<br>*Gender n (%)*: Female 79 (100)<br>*Ethnicity n (%)*: Latina, 79 (100)<br>Age: not recorded (NR)<br>*Diagnostic groups n (%)*: maternal depression 79 (100) | Phone call | None stated |
| Chen et al (23) | USA | CMHTs and outpatient services, VA service | To describe the implementation of TMH psychology services at a VA TMH hub | Descriptive study | **Service users** (N=252)<br>*Gender n (%)*: Male 226 (89.7), Female 26 (10.3),<br>*Ethnicity n (%)*: White non-Hispanic 182 (72.2), Black/African-American 40 (15.9), Hispanic/Latinx 4 (1.6), Asian/Pacific Islander 4 (1.6), Unknown 22 (8.7)<br>Age: mean 49.3 (range 21-88)<br>*Diagnostic groups n (%)*: depression 106 (42), trauma 77 (30.6), substance use 51 (20.2), anxiety 37 (14.7), sleep disorder 25 (9.9), bipolar 20 (7.9), adjustment disorder 20 (7.9), other/unknown 33 (13.1) | Video call | None stated |
| Felker et al (24) | USA | CMHTs and outpatient services | To describe the development, implementation, and evaluation of a TMH training programme, and consider whether such training programmes remain relevant given | Mixed-methods quality improvement project with 2- | **Staff** (n=100)<br>*Job title (%)*: Psychologist (37), social worker (22), not specified (19), psychiatrist (17), nurse (5)<br>Gender n (%) NR | Phone call | Reach-Effectiveness-Adoption-Implementation-Maintenance (RE-AIM) methodology <sup>a</sup> |
|  |  |  | the extent of TMH adoption in health care services | year follow-up survey | Age : NR |  |  |
| Hensel et al (25) | Canada | Crisis and emergency mental health services | To report on the perceived barriers surrounding the use of telepsychiatry for emergency assessments and an approach to overcoming those barriers for successful implementation of a programme to increase access to emergency psychiatric assessment | Survey to inform implementation with longitudinal outcome assessment | **Staff** (N=111)<br>*Job title n (%)*: Emergency physician 33 (30), psychiatric emergency nurse 14 (13), psychiatrist 33 (30), psychiatry resident 26 (23), physician assistant 2 (2), administrator 3 (3)<br>*Gender*: Male 64 (58), Female 44 (40), NR 3 (3) | Phone call | None stated |
| Lindsay et al (26) | USA | CMHTs and outpatient services | To report outcomes of implementation of a video telehealth evidence-based psychotherapy programme for post-traumatic stress disorder and pilot a facilitation strategy for implementation. | Implementation feasibility study | **Service users** (N=183)<br>*Gender n (%)*: NR<br>*Ethnicity n (%)*: NR<br>*Age*: NR<br>*Diagnostic groups n (%)*: NR | Video call | Promoting Action on Research Implementation in Health Services Framework with external facilitation as the primary strategy <sup>b</sup> |
| Lynch et al (27) | USA | CMHTs and outpatient services | To examine the service utilisation of a complex psychosis (CP) and non-CP cohort attending a largely group-based recovery-oriented behavioural health service before and after conversion to TMH | Retrospective cohort study and service evaluation | **Service users** (n=23 (CP participants); n=41 (non-CP))<br>*Gender n (%)*: Men 17 (74) CP, 20 (49) non-CP. Women 5 (22) CP, 17 (41) non-CP. Non-binary 1 (4) CP, 4 (10) non-CP.<br>*Ethnicity n (%)*: CP: White/Caucasian 20 (88) CP, 39 (95) non-CP. Black/African 1 (4) CP, 0 (0) non-CP. Hispanic/Latinx 1 (4) CP, 1 (2.5) non-CP. Asian 1 (4) CP, 1 (2.5) non-CP.<br>*Age*: Mean 32.6 CP, 26.1 non-CP.<br>*Diagnostic Groups n (%)*: CP 23 (35.9), non-CP 41 (64.1) | Video call | None stated |
| Lynch et al (28) | USA | CMHTs and outpatient services | To use mixed methods to understand the factors that contribute to successful telehealth conversion in a group-based recovery orientated service | Longitudinal cohort of service user utilisation outcomes and qualitative staff survey | **Staff** (N=6)<br>*Job title n (%)*: Practicing clinicians 6 (100)<br>*Gender n (%)*: NR<br>*Age*: NR<br>**Service users** (N=72, baseline demographics reported for n=60 participants)<br>*Gender n (%)*: Male 31 (51.7), Female 23 (38.3), Non-binary 6 (10)<br>*Ethnicity n (%)*: White/Caucasian 55 (91.7), Black/African American 1 (1.7), Hispanic/Latinx 2 (3.4), Asian 2 (3.4) | Video call | None stated |
|  |  |  |  |  | Age: Mean 28.1<br>Diagnostic groups n (%): psychotic disorder 15 (25), Autism spectrum disorder 15 (25), anxiety disorder 2 (3.4), affective disorder 28 (46.7) |  |  |
| Myers et al (29) | USA | CMHTs and outpatient services | To describe how VA Video Connect was implemented with a focus on the challenges of evidence-based practice delivery via TMH and VA Video Connect platforms | Prospective cohort and qualitative staff interview study | **Training:**<br>**Staff:** n=173 completed<br>*Job title (%)*: NR<br>*Gender n (%)* NR<br>*Age*: NR<br>**Qualitative interviews:**<br>**Staff:** n=8<br>*Job title (%)*: NR<br>*Gender n (%)* NR<br>*Age*: NR | Video call | Organisational champions <sup>c</sup> |
| Owens & Charles (30) | UK | CMHTs and outpatient services, Child and Adolescent Mental Health Services (CAMHS) | To test and refine a self-harming SMS text-messaging intervention (TeenTEXT) adapted for adolescents in CAMHS | Qualitative focus group and interview study | **Staff** (n=9 qualitative interviews, n=14 in one focus group)<br>*Job title (%)*: Interviews: Clinician 7 (77.8), Service manager 2 (22.2)<br>Focus group: CAMHS team members 14 (100)<br>*Gender n (%)* NR<br>*Age*: NR | Text messages | Normalisation Process Theory <sup>d</sup> |
| Puspitasari et al (31) | USA | CMHTs and outpatient services | To evaluate the feasibility and effectiveness of group-based transitional day programme for adults with transdiagnostic conditions at risk of psychiatric hospitalization that switched from in-person to video teletherapy during COVID-19 | Single arm non-randomised pilot study | **Service users** (n=76)<br>*Gender n (%)*: Male 10 (13), Female 65 (83), Transgender women 2 (3), Transgender men 1 (1).<br>*Ethnicity n (%)*: White 68 (90), African American 2 (3), Other 5 (7), NR 1 (1)<br>*Age: Mean* 36.55<br>*Diagnostic groups n (%)*: major depressive disorder 52 (68), bipolar disorder 6 (8), anxiety disorder 22 (29), personality disorder 13 (17), substance use disorder 6 (8), schizophrenia 2 (3) | Video call | None stated |
| Puspitasari et al (32) | USA | CMHTs and outpatient services, intensive outpatient programme | To describe the process for the rapid adoption and implementation of teletherapy in an intensive outpatient programme for adults with severe mental illness. | Pilot feasibility study | **Service users** (n=90)<br>*Gender n (%)*: NR<br>*Ethnicity n (%)*: NR<br>*Age*: NR<br>*Diagnostic groups n (%)*: NR | Video call, phone call | Implementation of teletherapy in the public sector model <sup>e</sup> |
| Sharma et al (33) | USA | Child psychiatry department in hospital | To investigate the implementation components involved in transitioning a comprehensive outpatient child and adolescent psychiatry programme to a home based TMH virtual clinic | Pilot Feasibility study | **Staff** (n=105)<br>*Job title n (%)*: clinical psychologist 51 (49), psychiatrist 34 (32), neurologist 1 (1), psychiatric nurse practitioner 7 (7) mental health therapist/behaviour analyst 12 (11)<br>*Gender n (%)*: NR<br>*Age*: NR | Video call, phone call | None stated |
| Taylor et al (34) | Australia | The Queensland Centre for Perinatal and Infant Mental Health | To investigate the importance of clinical facilitation for the implementation and sustainability of perinatal and infant mental health services | Qualitative staff interview study | **Staff** (n=14)<br>*Job title n (%)*: Medical officers, social workers, nurses, mental health clinicians, managers and health promotion workers (breakdown NR)<br>*Gender*: Male 3 (21), Female 11 (79)<br>*Age*: Range 26-62 | Video call, email | None stated |

<sup>a</sup>Glasgow, R. E., McKay, H. G., Piette, J. D., & Reynolds, K. D. (2001). The RE-AIM framework for evaluating interventions: what can it tell us about approaches to chronic illness management?. *Patient education and counseling*, 44(2), 119-127. <sup>b</sup>Rycroft-Malone, J. (2004). The PARIHS framework—a framework for guiding the implementation of evidence-based practice. *Journal of nursing care quality*, 19(4), 297-304. <sup>c</sup>Hendy, J., & Barlow, J. (2012). The role of the organizational champion in achieving health system change. *Social science & medicine*, 74(3), 348-355. <sup>d</sup>Murray, E., Treweek, S., Pope, C., MacFarlane, A., Ballini, L., Dowrick, C., ... & May, C. (2010). Normalisation process theory: a framework for developing, evaluating and implementing complex interventions. *BMC medicine*, 8(1), 1-11. <sup>e</sup>Muir, S. D., de Boer, K., Thomas, N., Seabrook, E., Nedeljkovic, M., & Meyer, D. (2020). Videoconferencing psychotherapy in the public sector: synthesis and model for implementation. *JMIR mental health*, 7(1), e14996.

### Quality of included studies

MMAT (18) quality appraisal was conducted for the 10 primary studies which aimed to answer a specific research question (as opposed to purely describing the implementation of TMH). Studies were generally of moderate to high quality, with seven studies meeting over 70% of quality criteria (average number of quality criteria met across all studies was 61%). Four descriptive studies were appraised using the AACODS checklist (19) and were also of high quality, with all studies meeting over 85% of quality criteria. As a result, all studies were given equal consideration during synthesis of results. A full breakdown of the results of the quality assessment is provided in Appendix 2.

### Evidence synthesis

Each type of implementation strategy in Eric’s taxonomy was reported in at least one study. The most commonly used strategy was ‘Train and educate stakeholders’, which was identified in nine studies, whilst the least used was ‘Utilise financial strategies’, which was only reported in one study (Hensel et al., 2020). The mean number of strategies used per study was 3.5, while the most common numbers of strategies used per study was 2 or 3. The implementation strategies reported by each study can be found in Table 2.

**Table 2:**
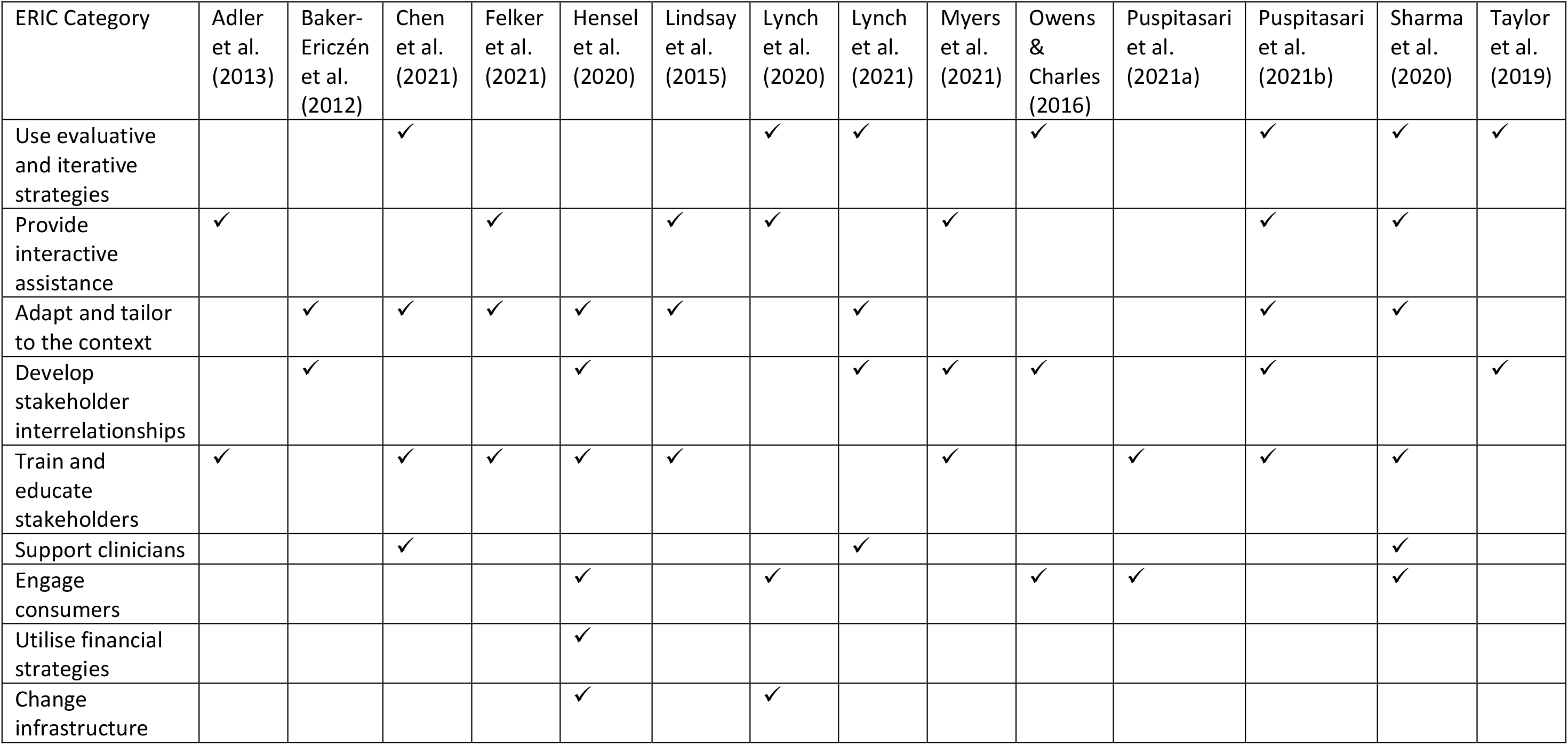
Implementation strategies used by each study

| ERIC Category | Adler et al. (2013) | Baker-Ericzén et al. (2012) | Chen et al. (2021) | Felker et al. (2021) | Hensel et al. (2020) | Lindsay et al. (2015) | Lynch et al. (2020) | Lynch et al. (2021) | Myers et al. (2021) | Owens & Charles (2016) | Puspitasari et al. (2021a) | Puspitasari et al. (2021b) | Sharma et al. (2020) | Taylor et al. (2019) |
| --- | --- | --- | --- | --- | --- | --- | --- | --- | --- | --- | --- | --- | --- | --- |
| Use evaluative and iterative strategies |  |  | ✓ |  |  |  | ✓ | ✓ |  | ✓ |  | ✓ | ✓ | ✓ |
| Provide interactive assistance | ✓ |  |  | ✓ |  | ✓ | ✓ |  | ✓ |  |  | ✓ | ✓ |  |
| Adapt and tailor to the context |  | ✓ | ✓ | ✓ | ✓ | ✓ |  | ✓ |  |  |  | ✓ | ✓ |  |
| Develop stakeholder interrelationships |  | ✓ |  |  | ✓ |  |  | ✓ | ✓ | ✓ |  | ✓ |  | ✓ |
| Train and educate stakeholders | ✓ |  | ✓ | ✓ | ✓ | ✓ |  |  | ✓ |  | ✓ | ✓ | ✓ |  |
| Support clinicians |  |  | ✓ |  |  |  |  | ✓ |  |  |  |  | ✓ |  |
| Engage consumers |  |  |  |  | ✓ |  | ✓ |  |  | ✓ | ✓ |  | ✓ |  |
| Utilise financial strategies |  |  |  |  | ✓ |  |  |  |  |  |  |  |  |  |
| Change infrastructure |  |  |  |  | ✓ |  | ✓ |  |  |  |  |  |  |  |

Most of the implementation strategies were reported as being associated with good outcomes for TMH implementation. However, some barriers to TMH implementation remained, including a lack of staff time, higher administrative burden, or a preference for face-to-face appointments (amongst either staff or service users). Five studies reported implementation outcomes resulting from the strategies used (Taylor 2019, Pupitsari 2021b, Myers 2021, Lyndsay 2015, and Felker 2021) while the remaining studies reported implementation outcomes only in relation to the TMH intervention itself. We had originally aimed to make specific links between strategies and outcomes, but as all but one study reported several implementation strategies in varying combinations, this was not possible. Instead, strategies and reported outcomes are outlined for each study below. Outcomes for each study are categorised according to the taxonomy of implementation outcomes (13). A summary of the strategies and outcomes for each study is presented in Table 3.

**Table 3:** Implementation strategies and outcomes for each study

| Author (year) | Implementation strategies used (ERIC Categories) | Implementation outcomes |
| --- | --- | --- |
| **Adler (2013)** | **Provide Interactive assistance**<br>Staff had monthly communication with therapists and met with clinical leaders every other month to discuss progress.<br>**Train and educate stakeholders**<br>Therapists completed online training and attended a video presentation by a psychotherapist with experience of TMH. | **Acceptability (Clinician views)**<br>Adopters reported that TMH was not as difficult or disruptive as they thought and were surprised by veteran acceptance of the approach. However, some clinicians reported little interest in using TMH.<br>**Feasibility**<br>Reported barriers included clinical demands, staff shortages, scheduling problems and equipment failures.<br>**Sustainability**<br>Two clinicians were offering TMH after 10 months. In many cases, clinical leaders had not acknowledged TMH as a priority. |
| **Baker-Ericzen (2012)** | **Adapt and Tailor to the Context**<br>The model used centrally located bilingual, bicultural Mexican-American mental health advisors to adapt to the cultural context and address barriers.<br>**Develop Stakeholder Interrelationships**<br>The model was designed to facilitate communication between primary care and mental health services using a mental health advisor. | **Acceptability (Service user and carer views)**<br>97% of mothers reported overall satisfaction with the intervention and 100% rated the quality of the mental health advisor as high.<br>**Fidelity**<br>Mental health advisors were trained using standardized procedures and followed a written treatment manual and study protocol. Fidelity ratings were 83%. |
| **Chen (2021)** | **Use Evaluative and Iterative Strategies**<br>Quality improvement data was gathered to allow rapid identification of problems and adjustments to be made.<br>**Adapt and Tailor to the Context**<br>Services were developed for TMH delivery based on a review of the literature and consultation with clinicians with previous experience of TMH.<br>**Train and Educate Stakeholders**<br>TMH was integrated into the existing Psychology training programmes, with the goal of offering TMH training to all existing Psychology training programmes within the next three years.<br>**Support Clinicians**<br>Five new psychologists were hired for the main hub, weekly calls were set up between spoke sites and hub staff to establish the services. One staff member served as the primary point of contact for each spoke. | **Adoption**<br>Within five months the service reached its preestablished productivity goals of 80 veteran encounters per month, per provider for the first year.<br>**Penetration**<br>From March 2017 to January 2018, 377 consults were received for TMH psychology services and 252 veterans engaged in TMH services. However, 32% did not receive treatment due to a variety of reasons, such as disengagement or discharge prior to services being offered. |
| **Felker (2021)** | **Provide Interactive Assistance**<br>Training courses and workshops to address the specific practical aspects of providing TMH. Clinicians were encouraged to engage in TMH with at least 2 patients and attend at least 10, 1-hour consultation calls to ask questions related to TMH (clinical or implementation issues).<br>**Adapt and Tailor to the Context**<br>Internal facilitators from each team provided consultation to external facilitators regarding the unique clinical and cultural aspects of their team (e.g. patients served, types of services provided, administrative needs, technological needs). External and internal facilitators tailored the TMH training programme to address clinic specific culture and barriers and meet unique clinic goals. | **Acceptability (Clinician views)\***<br>Following the training, 95% of providers agreed (n=42) or strongly agreed (n=35) that they were satisfied with the training provided.<br>**Adoption\***<br>Providers reported increased knowledge, skills and interest in TMH after training.<br>**Appropriateness\***<br>95% of providers agreed (n=50) or strongly agreed (n=28) that the amount of information covered was sufficient to begin using TMH. 76% of participants agreed (n=45) or strongly agreed (n=17) that they felt confident using TMH after receiving training.<br>**Feasibility**<br>Barriers identified included: lack of patient interest (45%), administrative burden (20%), preference for in- |
|  | **Train and Educate Stakeholders**<br>Clinical champions and team leads supported training and implementation of TMH. | person appointments (18%), concern about increased workload (11%), not completed all of the training components (6%), lack of supervisor support (4%), lack of provider interest (4%), and other reasons (4%). |
| Hensel (2020) | **Adapt and Tailor to the Context**<br>Initial survey of barriers allowed implementation to be tailored to the specific challenges identified by staff.<br>**Develop Stakeholder Interrelationships**<br>Worked with emergency departments to establish support staff available to assist with referral. Implementation leads were appointed at site and leadership at all levels were engaged in the programme. Clinical champions with TMH experience encouraged staff engagement.<br>**Train and Educate Stakeholders**<br>Education, anecdotes and evidence review from experienced providers. Initial training of a core group to develop expertise was conducted to build group confidence before engaging a larger cohort of providers. Training was offered to inexperienced providers.<br>**Engage Consumers**<br>Clear explanations were given to patients and families regarding the TMH programme.<br>**Utilize Financial Strategies**<br>Existing fee schedules were reviewed to support physicians and psychiatrists were salaried to avoid remuneration challenges. They also worked with regional authorities and hospitals to secure funding when needed.<br>**Change Infrastructure**<br>They worked with participating emergency departments to install dedicated equipment where possible or make arrangements regarding existing equipment. | **Adoption**<br>In the first year of operation, 243 assessments were completed.<br>Workload increased by 42% between the 6 months pre-programme and the second 6 months of programme operation. There was a 2% increase in presentations at the hub, and some increase in workload from the spokes which saw declines in on-site support and an 8% increase in total mental health and addiction presentations. The percentage of transfers avoided increased from 0% pre-programme to 65% in December 2018. |
| Lindsay (2015) | **Provide Interactive Assistance**<br>Technical support was provided through weekly consultation calls with a facilitator to discuss technical and logistical issues specific to the delivery of TMH.<br>**Adapt and Tailor to the Context**<br>Site-specific implementation plans were tailored to unique needs of the site including needs of stakeholders<br>**Train and Educate Stakeholders**<br>Intensive training in evidence-based practice for PTSD was provided to providers including an experientially orientated 2-3-day workshop and weekly consultations with experts. | **Acceptability (Clinician views)\***<br>Therapists reported a high degree of satisfaction and rated the external facilitation model as very helpful in their efforts to implement video telehealth (6.67 out of 7), viewing the regular facilitation calls as very important to establishing video telehealth services.<br>**Penetration**<br>Compared to baseline, participating sites averaged a 6.5-fold increase in psychotherapy sessions conducted via TMH, whereas non-participating sites averaged a 1.7-fold increase. |
| Lynch (2020) | **Use Evaluative and Iterative Strategies**<br>In response to reports of problems with maintaining attention in virtual sessions, clinicians problem solved with clients to minimise distractions, used screen sharing features and interactive activities, and provided additional brief breaks when needed.<br>**Provide Interactive assistance**<br>Virtual training on the features and functionality of telehealth platforms were provided to staff.<br>**Support Clinicians**<br>Factors to support and capture work from home productivity were considered for staff.<br>**Engage Consumers**<br>Individualized instruction about telehealth platforms were provided to service users as needed. | **Adoption**<br>TMH acceptance rates indicated that 90% (n=18) of the 20 patients enrolled at the time of conversion agreed to TMH sessions within ten days of the service transition and maintained their specific treatment plans virtually. An additional five service users began using TMH after the start of the study. There were no significant differences in attendance rates before conversation to TMH, and no differences in acceptance between the TMH and non-TMH group.<br>**Feasibility**<br>Following conversion to TMH, participants and clinicians sought to maintain individualized treatment plans and group schedules whenever possible, which may have contributed to the high acceptance rates and unchanged service utilization. |
| Lynch (2021) | **Use Evaluative and Iterative Strategies**<br>The service responded to challenges identified by staff with new implementation strategies.<br>**Adapt and Tailor to the Context**<br>Group session material was adapted to be engaging on virtual platforms.<br>**Develop stakeholder interrelations**<br>In addition to formal systems that were put in place to ensure consistent communication (e.g., end-of-day email debriefs), staff had increased support from supervisors to facilitate both client care coordination and opportunities for staff to “support each other as individuals.”<br>**Engage Consumers**<br>Through a collaborative approach some service users who were challenged by TMH helped the team to come up with web etiquette guidelines for other service users.<br>**Change Infrastructure**<br>The proactive culture at the clinic helped rapid transition to TMH and aided continuity of care. Resources, workflows and infrastructure were developed in anticipation of regulatory change, rather than in response. | **Acceptability (Clinician views)**<br>Though staff perceived the shift to TMH as slightly more challenging for themselves than for clients, they reported learning to navigate the technology and virtual interaction fairly quickly. However, TMH negatively impacted staff’s ability to communicate with each other, due to the lack of informal contacts. ‘Zoom fatigue’ and exhaustion were also reported by staff.<br>**Acceptability (Service user and carer views)**<br>All respondents who completed the questionnaire (n=18) provided a score >23, suggesting satisfaction with the TMH services. However, 78% of respondents stated that they would still prefer in-person sessions if there were no health risks. Only 50% reported feeling that TMH was as good as in-person sessions.<br>**Adoption**<br>93% of service users enrolled at the time of conversion agreed to maintain their specific treatment plans virtually. 7% opted out. Session attendance did not significantly differ over time or between in-person and TMH formats. The mean no show/cancellation rate was 37% less at 13-18 weeks after implementation of TMH compared to in-person (B= -.47, p < 0.05).<br>**Appropriateness**<br>TMH was deemed appropriate because of its increased flexibility to adapt scheduling to client capacity for engagement, e.g. offering shorter, more frequent breaks, or reducing session duration but increasing frequency. However, staff raised concerns that for some service users, long-term TMH utilization may hinder recovery, as the routine and engagement associated with traveling to a clinic may enhance treatment investment and pro-health behaviours.<br>**Feasibility**<br>Staff found TMH more challenging for clients who had technology or gaming addictions, or symptoms associated with attention deficit hyperactivity disorder or autism.<br>**Fidelity**<br>Staff noted that group dynamics in virtual sessions were largely positive and similar to in-person sessions, with clients interacting with one another and not responding solely to the group leader. |
| Myers (2021) | **Provide Interactive Assistance**<br>TMH champions assisted with enrolment into the TMH system, procurement of equipment and completion of a systems check (e.g., test calls, quality check of audio and visual issues).<br>**Develop Stakeholder Interrelationships**<br>Site champions (with previous experience or trained for leadership roles) were utilised to support implementation.<br>**Train and Educate Stakeholders**<br>The TMH champions assisted with mandatory training of policy and procedures, and with selection criteria for determining appropriateness of treatment via TMH. | **Adoption**<br>The site failed to address lack of internet or phone access for service users, which affected implementation. However, use of TMH was increased by 42%.<br>**Appropriateness**<br>TMH was considered appropriate other than for suicidal or psychotic individuals. Lack of appropriateness for these service users, however, limited the ability to provide crisis support.<br>**Feasibility**<br>Providers reported concerns about the feasibility of TMH: 1) it reduced their ability to respond to emergencies (e.g., responding to suicidal patients); 2) it may not be feasible for some veterans considered “too high risk” or unstable; 3) some veterans were not respecting therapeutic boundaries (e.g., trying to engage in treatment sessions while driving); 4) too much time was lost attending to technical issues; and 5) difficulty in delivering measurement-based care.<br>**Implementation Cost\***<br>The main cost was time-related (the role of site champion was unpaid).<br>**Sustainability**<br>Sustainability of TMH may vary by site, depending on organisational constraints (administration, other role commitments which may inhibit implementation and ongoing support).<br>It is unclear if all providers should be “telehealth generalists” or if TMH should be a speciality. |
| Owens (2016) | **Use Evaluative and Iterative Strategies**<br>Clinicians and service users worked closely with the research team and software developers through a series of three iterations or feedback loops to optimise the intervention and assess whether it was sufficiently likely to normalise to be worth evaluating in a full trial.<br>**Develop stakeholder interrelations**<br>Three clinicians in each team supported and mentored each other for the duration of the study and cascaded knowledge through the team, influencing others to adopt the intervention. | **Acceptability (Clinician views)**<br>Clinicians saw it as a potentially valuable tool to help young people manage their self-harming behaviour.<br>**Adoption**<br>The most significant barrier to adoption was the need for buy-in at management levels and the time it took to obtain this.<br>**Feasibility**<br>CAMHS teams reported being under very high pressure which negatively affected their ability to be involved in new projects.<br>**Appropriateness**<br>In the context of very heavy caseloads, high stress levels and exhaustion, the effort involved in mastering a new technology and incorporating it into everyday practice was perceived to be too much by clinicians. Although some reported that they were using apps of various kinds with their clients, others appeared to be resistant to technological interventions. Nearly all informants believed that CAMHS was not the ideal delivery setting as clinicians see only the most acute and complex cases and duration of contact with CAMHS is typically short. |
| Puspitasari (2021a) | **Train and educate stakeholders**<br>Counsellors attended weekly consultation meetings facilitated by a clinical psychologist to ensure treatment adherence and fidelity. All disciplines attended daily meetings to discuss safety management and patient progress.<br>**Engage Consumers**<br>Service users who were accepted into the programme received assistance from programme staff and information technology support staff to prepare for the first TMH session. Each group was led by two counsellors: one as the primary facilitator leading the presentation and group discussion, the other assisting patients with any technological issues. | **Adoption**<br>The completion rate of the programme was 70/76. This completion rate was higher than typical completion rates for psychiatric Intensive outpatient or partial hospitalization programmes.<br>**Feasibility**<br>Zoom features (including chat, whiteboard, shared screen and waiting room) improved feasibility. It was also feasible to conduct psychotherapy experiential exercises via videoconferencing, e.g., performing guided group mindfulness exercises, completing psychotherapy forms, and watching psychotherapy skills videos. |
| Puspitasari (2021b) | **Use Evaluative and Iterative Strategies**<br>A staged implementation strategy was used where the TMH group intervention was first piloted in one site, which indicated readiness for TMH implementation, openness among clinicians and availability of resources. Challenges faced during the TMH rollout were informally assessed and communicated to team members for efficient problem solving.<br>**Provide Interactive Assistance**<br>A multi-disciplinary TMH committee coordinated the change to TMH and ensured clinicians had access to necessary technology. All clinicians had 24/7 access to the IT help desk for additional support. An operations manager coordinated the preparation, adoption, and implementation phase. This individual was responsible for managing the workflow and engaging other stakeholders within and outside of the department to ensure a smooth transition to teletherapy. Quick reference guides were also created for clinicians to help them adapt to TMH.<br>**Adapt and Tailor to the Context**<br>The committee met twice weekly for the first month during the most rapid phase of implementation to review, update and expand upon existing training resources, guidelines, and policies. Due to the closing of many behavioural health services in the surrounding area and increased need for intensive outpatient care, the service expanded capacity and added an additional intervention for patients suffering with mood and anxiety disorders.<br>**Develop Stakeholder Interrelationships**<br>TMH champions were identified (including directors, an operations manager, committee | **Adoption**<br>Education, training, and ongoing supervision were of particular importance at the start of teletherapy implementation to support clinicians' successful engagement with the technology, as well as to establish an effective practice for virtual therapy.<br>**Feasibility**<br>Data on patient attrition indicated that TMH was feasible to assure patient retention, since many service users completed the programme and the average number of sessions attended was high.<br>**Penetration**<br>A plan was established by the pilot site to initiate full implementation following the pilot. |
|  | members, IT specialists, and several clinicians with TMH experience or enthusiasm). These champions were fully integrated into the team to provide adequate support for its other members. Daily virtual meetings attended by all staff allowed discussion of patient progress and issues.<br>**Train and Educate Stakeholders**<br>Education, training, and ongoing supervision were integral implementation strategies prior to TMH adoption. |  |
| Sharma (2020) | **Use Evaluative and Iterative Strategies**<br>Pilot tests were conducted with three small groups of parents, with satisfaction surveys resulting in a change of platform.<br>**Provide Interactive Assistance**<br>A brief technical guide was provided to all clinicians after group TMH training sessions to assist in their subsequent TMH clinics. A “cheat sheet” was developed to help the clinician guide families through the process of setting up their home systems and responding to the e-invite for a TMH session.<br>**Adapt and Tailor to the Context**<br>Each day the faculty analysed and adapted to latest government rules regarding stay-at-home mandates and patient and staff needs.<br>**Train and Educate Stakeholders**<br>Videoconferencing training sessions were run to quickly train staff on the online platform and clinical aspects of TMH.<br>**Engage Consumers**<br>If a family was not able to participate in TMH due to lack of internet access, then a phone appointment was offered to ensure equity. | **Adoption**<br>Failure of the outpatient videoconferencing platform delayed full home-based TMH adoption.<br>**Feasibility**<br>This study demonstrates the feasibility of rapidly building upon an existing telemedicine infrastructure to train a large group of multidisciplinary providers to deliver urgent home-based TMH services. However, the key message is that even with a well-established telemedicine infrastructure, programmes must expect to encounter serious challenges during crises. Planning for the next crisis should start now.<br>**Implementation Cost**<br>Funding fell dramatically after transitioning patients from clinic to home. Interim phone appointments while awaiting full implementation of TMH services yielded less revenue per appointment than in-clinic or TMH appointments, although required the same amount of time and almost the same level of documentation by the faculty.<br>**Penetration**<br>After 1 month, TMH was offered to all established outpatients for individual visits and the clinic started a trial process for enrolling new patients. Continued work on expanding TeleGroups occurred. Only the crisis clinic continued a regular in-clinic presence.<br>**Sustainability**<br>The faculty’s relatively rapid but complex development of clinic-wide home-based TMH and TeleGroups was reported to help to advance and increase access to psychiatric care. Authors argued that in the future, home-based TMH may help overcome barriers to treatment such as distance, transportation and scheduling. |
| Taylor (2019) | **Use evaluative and iterative strategies**<br>A pilot project established the efficacy of the intervention in improving the skills and knowledge of local health service providers but identified a need for additional clinical support in specialist areas. This was therefore integrated into the model.<br>**Develop Stakeholder Interrelationships**<br>As a result of the pilot project, General practitioners (GPs), mental health professionals and other service providers were offered access to secondary consultations with perinatal and infant psychiatrists. The service also employed a clinical facilitator who was responsible for service promotion, site visits, staff education and training, co-ordinating case conferences and video consultations. | **Acceptability (Clinician views)**<br>Mental health workers who had used TMH were unanimously complimentary about the service, reporting that it allowed expert input into care planning, reduced professional isolation, upskilled remote workers and provided a sense of security for remote care providers.<br>**Appropriateness**<br>The study showed that TMH can help address unmet need for specialist mental health services in regional, rural and remote areas.<br>**Sustainability\***<br>Clinical facilitation is likely to be more important in intermittent compared with high-volume services where regular clinics can make TMH more visible. Ongoing facilitation is necessary for the sustainability of TMH services due to intermittent demand and local impediments, such as fragmentation of service providers and transiency of the workforce. |
| * outcomes of the strategy to improve implementation of the TMH intervention (vs outcomes of the intervention itself) |  |  |

#### Adler et al. (2013)

This study used two main categories of implementation strategies as part of a pilot project to improve the delivery of TMH in a Veterans Affairs (VA) service: ‘provide interactive assistance’ and ‘train and educate stakeholders’. Staff had monthly communication with therapists and met with clinical leaders every other month to discuss progress. Therapists completed online training and attended a video presentation by a psychotherapist with experience of TMH. Following the use of these strategies, the authors reported on three types of outcome relating to the TMH intervention: acceptability, feasibility, and sustainability. Acceptability outcomes varied between clinicians, with some reporting that TMH was not as difficult or disruptive as they had expected it to be, and that veteran acceptance of the approach surprised them. However, others reported little interest in conducting TMH. TMH was not viewed as feasible by all, with identified barriers to implementation including clinical demands, staff shortages, scheduling problems and equipment failures. TMH was not well sustained, as only two clinicians were offering TMH after 10 months. In many cases, clinical leaders had not acknowledged TMH as a priority.

#### Baker-Ericzén et al. (2012)

This study used two types of implementation strategies in adopting a culturally adapted telemedicine intervention for Latina women with maternal depression. The first strategy used was ‘adapt and tailor to the context’, as the model used centrally located bilingual, bicultural Mexican American mental health advisors to adapt to the cultural context and address barriers. The second strategy was ‘develop stakeholder interrelationships’, as the model was also designed to facilitate communication between primary care and mental health services using a mental health advisor. These strategies were associated with high acceptability of the TMH intervention, as 97% of mothers reported overall satisfaction with the intervention and 100% rated the quality of the mental health advisor as high. Fidelity ratings of the intervention were also high, with a score of 83%.

#### Chen et al. (2021)

The authors described the implementation of TMH psychology services at a VA TMH hub. Four categories of implementation strategy were used: ‘use evaluative and iterative strategies’, ‘adapt and tailor to the context’, ‘train and educate stakeholders’, and ‘support clinicians’. Quality improvement data was gathered to allow rapid identification of problems and adjustments to be made. Services were developed for TMH delivery based on a review of the literature and consultation with clinicians with previous experience of TMH. TMH was integrated into the existing psychology training programmes, with the goal of offering TMH training to all existing psychology training programmes within the next three years. Five new psychologists were hired for the main hub, weekly calls were set up between spoke sites and hub staff to establish the services. One staff member served as the primary point of contact for each spoke. These four strategies were associated with moderate to high adoption and penetration outcomes for the TMH intervention, as within five months the service reached its pre-established productivity goals of 80 veteran encounters per month, per provider for the first year. In the first nine months, from March 2017 to January 2018, 377 consults were received for TMH psychology services and 252 veterans engaged in TMH services. However, 32% did not receive treatment due to a variety of reasons, such as disengagement or discharge prior to TMH services being offered.

#### Felker et al. (2021)

This study described the development, implementation, and evaluation of a TMH training programme. Three strategies were used, the first of which was ‘provide interactive assistance’, in which training courses and workshops were conducted to address the specific practical aspects of providing TMH. Clinicians were encouraged to engage in TMH with at least two patients and attend at least ten one-hour consultation calls to ask questions related to TMH. Secondly, they used ‘adapt and tailor to the context’, in which internal facilitators from each team provided consultation to external facilitators regarding the unique clinical and cultural aspects of their team (e.g. patients served, types of services provided, administrative needs, technological needs). External and internal facilitators tailored the TMH training programme to address clinic specific culture and barriers and meet unique clinic goals. Finally, they used ‘train and educate stakeholders’, in which clinical champions and team leads supported training and implementation of TMH. Three outcomes relating to these strategies were reported. Clinicians viewed TMH as acceptable, as following the training, 95% of providers agreed (n=42) or strongly agreed (n=35) that they were satisfied with the training provided. Other implementation outcomes of these strategies were adoption, as providers reported increased knowledge, skills and interest in TMH after training, and appropriateness, as 95% of providers agreed (n=50) or strongly agreed (n=28) that the amount of information covered was sufficient to begin using TMH. 76% of participants agreed (n=45) or strongly agreed (n=17) that they felt confident using TMH after receiving training. Feasibility outcomes of the TMH intervention itself were also reported, with identified barriers to successful implementation following the above strategies including: lack of patient interest (45%), administrative burden (20%), preference for in-person appointments (18%), concern about increased workload (11%), not completed all of the training components (6%), lack of supervisor support (4%), lack of provider interest (4%), and other reasons (4%).

#### Hensel et al. (2020)

This study reported on the barriers surrounding the use of telepsychiatry for emergency assessments and described an approach to overcoming those barriers for successful implementation of a programme to increase access to emergency psychiatric assessment. This study employed six different implementation strategies: ‘adapt and tailor to the context’ (an initial survey of barriers allowed the implementation to be tailored to address these), ‘develop stakeholder interrelationships’ (by using clinical champions and encouraging staff engagement), ‘train and educate stakeholders’, ‘engage consumers’ (clear explanations were given to patients and families regarding TMH), ‘utilize financial strategies’ (secured funding and reviewed the fee schedule), and ‘change infrastructure’ (installed dedicated equipment and made arrangements regarding existing equipment). Following these strategies, adoption (of the TMH intervention) implementation outcomes were reported, indicating successful adoption of TMH (for example, 243 assessments were completed and the percentage of transfers to other hospitals that were avoided increased from 0% pre-programme to 65% in December 2018).

#### Lindsay et al. (2015)

This study reported outcomes of implementation of a video telehealth evidence-based psychotherapy programme for post-traumatic stress disorder and the results of a pilot facilitation strategy for implementation. Three implementation strategies were reported: ‘provide interactive assistance’, ‘adapt and tailor to the context’, and ‘train and educate stakeholders’. Technical support was provided through weekly consultation calls with a facilitator to discuss issues specific to the delivery of TMH. Site-specific implementation plans were tailored to the unique needs of the site, including needs of stakeholders. Intensive training in evidence-based practice for PTSD was also given to providers. One outcome directly related to these three strategies was reported, alongside one related to the TMH intervention. Clinicians reported a high degree of satisfaction and viewed the external facilitation model as very helpful in implementing video consultations (6.67 out of 7). They also found the regular facilitation calls to be very important in establishing video telehealth services. Penetration of the intervention was also reported: compared to baseline, participating sites averaged a 6.5-fold increase in psychotherapy sessions conducted via TMH, whereas non-participating sites only averaged a 1.7-fold increase.

#### Lynch et al. (2020)

This study used four different implementation strategies to examine the service utilisation of a complex psychosis (CP) and non-CP cohort attending a largely group-based recovery-oriented behavioural health service before and after conversion to TMH: ‘use evaluative and iterative strategies’, ‘provide interactive assistance’, ‘support clinicians’ and ‘engage consumers’. In response to reports of problems with maintaining attention in virtual sessions, clinicians problem solved with clients to minimise distractions, used screen sharing features and interactive activities and provided additional brief breaks when needed. Virtual training on the features and functionality of telehealth platforms was provided to staff, factors to support and capture work from home productivity were considered for staff and individualized instruction regarding telehealth platforms was provided to service users as needed. Adoption and feasibility outcomes for the TMH intervention were good following the use of these strategies: for example, 90% of patients who were enrolled in the service agreed to telehealth sessions, and individualized treatment plans and group schedules were maintained following conversion to TMH.

#### Lynch et al. (2021)

This study explored factors which influenced successful conversion to telehealth in a group-based recovery orientated service. It reports the use of five different implementation strategies: ‘use evaluative and iterative strategies’, where the service responded to challenges identified by staff; ‘adapt and tailor to the context’, where group session material was adapted to be engaging on virtual platforms; ‘develop stakeholder interrelations’, with increased communication and support for staff; ‘engage consumers’, where, via a collaborative approach, some service users who found the use of TMH challenging helped the team develop web etiquette guidelines for other service users; and ‘change infrastructure’, facilitated by a “proactive culture at the clinic” when implementing TMH. Several implementation outcomes for the TMH intervention were reported following the use of these strategies: findings indicated high acceptability and adoption, TMH was viewed as appropriate for this patient group and fidelity of the group intervention was high. However, TMH was not viewed as feasible for everyone, as staff found it more challenging for clients who had technology or gaming addictions, or symptoms associated with attention deficit hyperactivity disorder or autism.

#### Myers et al. (2020)

This study used three strategies (‘provide interactive assistance’, ‘develop stakeholder interrelationships’ and ‘train and educate stakeholders’) during the implementation of VA Video Connect (a TMH platform). TMH champions assisted with enrolment into the system, procurement of equipment and completion of systems checks (for example, test calls, quality checks of audio and visual issues). In addition, site champions with previous experience supported implementation by assisting with mandatory training on policy and procedures and with developing selection criteria for determining appropriateness of treatment via TMH. Following the use of these these strategies increased adoption of TMH was reported, with usage increasing by 42%. TMH was considered largely appropriate other than for suicidal or psychotic individuals, as providers expressed concerns about managing risk. There were also concerns around the feasibility of TMH for some service users, as it was deemed less suitable for those at ‘high risk’, those in crisis, and those without telephone or internet access. In terms of sustainability, the study concluded that this varied across sites depending on organisational constraints, such as administration time or other role commitments. Implementation cost was reported in relation to the use of site champions as a strategy, with the main cost being an increase in staff workload, as no additional funding was available for the role of site champion.

#### Owens & Charles (2016)

This study used two implementation strategies in investigating the use of a self-harming SMS text-messaging intervention (TeenTEXT) adapted for adolescents in child and adolescent mental health services (CAMHS). The first strategy was ‘use evaluative and iterative strategies’, in which clinicians and service users worked closely with the research team and software developers through a series of three iterations or feedback loops to optimise the intervention. The second strategy was to ‘develop stakeholder interrelations’, in which three clinicians in each team supported and mentored each other for the duration of the study and cascaded knowledge through the team, influencing others to adopt the intervention. Implementation outcomes following the use of these strategies were mixed: although clinicians viewed the intervention as acceptable, barriers to adoption and feasibility of the TMH intervention were identified as CAMHS teams reported being under high pressure before implementation, which limited their time to learn the intervention. The intervention was not viewed as appropriate for a CAMHS setting as clinicians saw only the most acute and complex cases and duration of contact with CAMHS is typically short.

#### Puspitasari et al. (2021a)

This study used two strategies to promote implementation of a group-based transitional day programme for adults with transdiagnostic conditions at risk of psychiatric hospitalization. Firstly, the strategy ‘train and educate stakeholders’ was used, with counsellors attending weekly consultation meetings facilitated by a clinical psychologist to ensure treatment adherence and fidelity. Secondly, they employed the strategy ‘engage consumers’, in which service users who were accepted into the programme received assistance from programme staff and information technology support staff to prepare for the first TMH session. An additional counsellor was on hand to assist patients with any technological issues during group sessions. Two implementation outcomes relating to the TMH intervention were investigated following the deployment of these strategies: adoption and feasibility. The completion rate of the programme was 70/76, which was higher than typical completion rates for psychiatric intensive outpatient or partial hospitalization programmes. The use of several Zoom features, including chat, whiteboard, screen sharing and waiting room, improved feasibility. It was also feasible to conduct psychotherapy experiential exercises, such as mindfulness, via videoconferencing.

#### Puspitasari et al. (2021b)

This study reported on the use of five implementation strategies for the rapid adoption and implementation of teletherapy due to the COVID-19 pandemic in an intensive outpatient programme for adults with severe mental illness. These were: ‘use evaluative and iterative strategies’, ‘provide interactive assistance’, ‘adapt and tailor to the context’, ‘develop stakeholder interrelationships’, and ‘train and educate stakeholders’. The overarching approach was to involve a multi-disciplinary TMH committee who coordinated the change to TMH by providing training, ensuring clinicians had access to necessary technology and IT support, reviewing and expanding guidelines and policies, identifying TMH champions and providing ongoing support and supervision. Following the deployment of these strategies three outcomes relating to the TMH intervention were measured: adoption, feasibility and penetration. The study identified education, training, and ongoing supervision as being particularly important in increasing adoption and engaging clinicians. Data on patient attrition indicated that TMH is feasible to assure patient retention, since many service users completed the programme and the average number of sessions attended was high. A plan had also been established by the pilot site to initiate full implementation following pilot implementation, indicating high penetration.

#### Sharma et al. (2020)

This study used five implementation strategies in investigating the implementation components involved in transitioning a comprehensive outpatient child and adolescent psychiatry programme to a home-based TMH virtual clinic. These were: ‘use evaluative and iterative strategies’, ‘provide interactive assistance’, ‘adapt and tailor to the context’, ‘train and educate stakeholders’, and ‘engage consumers’. For example, pilot tests were conducted with three small groups of parents, technical guidance was provided to all clinicians after group TMH training sessions, and a “cheat sheet” was developed for families to help them access TMH. Each day the clinic analysed and adapted to the latest government rules regarding stay-at-home mandates, and to patient and staff needs. If a family was not able to participate in the intervention due to lack of internet access, then a phone appointment was offered to ensure equity. These strategies resulted in five outcomes relating to the TMH intervention. Adoption of TMH was delayed by failures of the videoconferencing platform, however penetration of telemental heath was overall successful, as by April 10^th^ 2020, all established outpatients were offered remote appointments. Findings also indicated that these strategies meant it was feasible to rapidly expand the existing telehealth infrastructure during an emergency. In terms of implementation cost, the service reported that less funding was generated from interim phone appointments (before proper TMH could take place) than from face-to-face or TMH appointments. The study authors viewed TMH as sustainable and as having the potential to help overcome barriers to treatment, such as distance, transportation and scheduling.

#### Taylor et al. (2019)

This study used two strategies - ‘use evaluative and iterative strategies’ and ‘develop stakeholder interrelationships’ - to investigate the importance of clinical facilitation for the implementation and sustainability of TMH in perinatal and infant mental health services. Firstly, a pilot project established the efficacy of the intervention in improving the skills and knowledge of local health service providers but identified a need for additional clinical support in specialist areas, which was therefore integrated into the model. As a result of a pilot project, General Practitioners (GPs), mental health professionals and other service providers were offered access to secondary care consultations with perinatal and infant psychiatrists. The service also employed a clinical facilitator who was responsible for service promotion, site visits, staff education and training, co-ordinating case conferences and video consultations. Two outcomes relating to the TMH intervention were reported following the deployment of these implementation strategies: acceptability and appropriateness. All mental health workers who had used TMH evaluated it positively, reporting that it allowed expert input into care planning, reduced professional isolation, upskilled remote workers and provided a sense of security for remote care providers. Regarding appropriateness, the study showed that TMH can help address unmet need for specialist mental health services in regional, rural and remote areas. In terms of the implementation strategies themselves, the study concluded that ongoing clinical facilitation is necessary for the sustainability of TMH services due to intermittent demand and local impediments, such as fragmentation of service providers and transiency of the workforce, which can make continued unsupported use of new technology challenging.

## Discussion

### Summary of findings

In this study, we have reviewed literature pre- and post-COVID-19 on strategies used to improve implementation of TMH and associated implementation outcomes. We identified as meeting our inclusion criteria fourteen studies, conducted across five countries. Both staff and service user views and outcomes were represented in these studies. Results indicated that using a combination of different planned implementation strategies could be associated with successful implementation of TMH, although the methodologies of most studies were such that firm causal conclusions were difficult to draw. Whilst we had originally planned to explore links between individual types of implementation strategy and implementation outcomes directly, we were unable to isolate the effects of specific mechanisms as all studies reported outcomes of initiatives that combined multiple implementation strategies. We are, however, able to propose some tentative conclusions based on the synthesis of findings from studies which reported outcomes of these strategies. Ongoing support and facilitation, for example, through either technical assistance or ongoing consultation, was common and tended to be strongly linked to successful implementation. Providing initial training and the use of ‘digital champions’ to model best practice, also benefited implementation of TMH. Other recent studies further support and supplement these conclusions. In our recent rapid realist review of TMH (35), we found that providing staff with training on the use of technology to deliver mental health services, a strategy reported by several studies in the current review, was reported to increase confidence in and uptake of TMH. The rapid realist review also found that adapting the use of TMH to take into account service user preferences was beneficial in removing barriers to accessing TMH (35) (see Box 1 for further discussion).

Research indicates that ERIC strategies are considered suitable to influence different implementation outcomes (36), but there is currently little consensus or evidence regarding which strategies affect specific outcomes. Furthermore, Powell and colleagues (37) argue that implementation strategies should be tailored to the circumstances and context of each change project, as they may be more likely to result in changes to practice. They suggest four different methods to identify appropriate strategies: concept mapping, group model building, conjoint analysis, and intervention mapping. Essentially, this means that the appropriateness and effectiveness of an implementation strategy or implementation support ‘bundle’ may well depend on the context in which a clinical intervention or service delivery mechanism, such as TMH, are introduced to. It logically follows that the same implementation strategy may be very well suited to one context, but redundant in another if it fails to address specific barriers to implementation. Hence selection of implementation strategies should be tailored to the local context of application. Whilst some studies included in this review evaluated the barriers and facilitators to TMH before using implementation strategies to address these, not all papers reported taking this approach in a systematic manner.

### Implications for future research

As all studies reported using multiple implementation strategies, we were unable to compare the effectiveness of specific strategies. This could be addressed by future research, for example, cluster randomised controlled trials to formally compare the effectiveness of different implementation strategies linked to specific outcomes, although it would also be helpful to evaluate a theoretically informed approach to selecting a bundle of implementation strategies to fit a particular context, using more robust study designs. Inclusion of a control group in future evaluations of the implementation of TMH is critical: without this we will remain unable to establish causal links between the presence or absence of a strategy to support implementation, or bundle thereof, and success of implementation(38) offers an illustration of a clustered randomised evaluation of different interventions to support the implementation of four evidenced-based psychosis treatments. Within this study, what was randomised was not the clinical therapies, but rather the level of implementation support, which was limited in the control arm (provision of treatment manuals) and substantially enhanced in the intervention arm to include toolkits, training, implementation facilitation, and data-based feedback. The primary endpoint was fidelity of treatment delivery, and the trial concluded that the implementation ‘bundle’ was successful in enhancing fidelity across all four studied treatments for psychosis. Similarly designed studies in the context of TMH provision will significantly expand our knowledge regarding how best to deliver it sustainably and effectively.

Beyond controlled evaluations, our understanding of the relative effectiveness and suitability of strategies to support implementation of TMH across different settings can be further enhanced if future observational studies offer a detailed description of local (or wider, as appropriate) settings in which TMH is offered; and a well-articulated rationale for the selection of strategies (such as those we summarised above) to support implementation – such that subsequent evidence syntheses can offer a better articulation of which strategies may be better suited to which contexts. We further propose that selection of implementation support interventions should be based on a description and mapping of the barriers and drivers an implementation effort is likely to face, for example carried out using one of the methods proposed by Powell et al (36).

It is also important to note that most studies identified in this review were conducted in the United States. There is therefore a need to replicate these findings and conduct further research in other countries with different healthcare structures and funding models to generalise the findings from this research.

### Implications for practice

Our review identified a range of potential implementation strategies to be deployed to improve TMH implementation in routine settings, with evaluations spanning a full range of types of implementation strategy. Although we were unable to identify causal links between implementation strategies and outcomes, findings across the included studies suggest that when implementing TMH, service planners should consider a multi-component implementation strategy. This strategy should be tailored to the local context and designed to address any pre-identified barriers. It is likely that staff training and facilitation support are key factors in the success of implementing TMH.

### Strengths and limitations

A strength of our review is that it focused only on studies which implemented TMH as part of their routine service (i.e., not just in a trial), which means findings are more likely to generalise to ‘real world’ settings. This review also integrated data from studies conducted before the COVID-19 pandemic with those conducted during the pandemic, which enables findings to be used to inform future models of service development. Included studies were mostly of moderate to high quality, however only five interventions were informed by an established implementation framework. Limitations of this research should be acknowledged. Firstly, as noted earlier, the high heterogeneity of strategies and outcomes reported across studies makes it hard to reach firm judgements about which strategies are linked with effective implementation of TMH. As all studies reported the inclusion of multiple strategies, we were unable to draw conclusions regarding the active ingredients of specific strategies. Secondly, researchers were not blinded to the results of screening and quality assessment during double screening. This was due to the short timeframe in which the review was conducted in order to make its results relevant to the current service context.

#### Box 1 Lived experience commentary by Beverly Chipp & Karen Machin, members of the NIHR Mental Health Policy Unit’s Lived Experience Working Group.

This systematic review reveals the lack of knowledge on how best to implement telemental health (TMH). The included studies refer to a range of settings, dates, TMH applications and patient groups making it difficult to draw a single conclusion which might work across them. Participants’ age likely affects affinity for technology (not recorded in 8 studies), and other demographic and cultural factors may impact on access. Implementation strategies might vary across healthcare systems internationally, and the pandemic impacted on available resources and funding streams which may have influenced procurement choices.

The underlying assumption is that TMH is beneficial. New technology is generally viewed as progress, but in health and social care the most important consideration should be human relationships, both with patients and between staff. Technologies may disturb these relationships and the full implications upon both workforce and healthcare are yet to be considered. For example, service users emphasise that choices are essential, including the option of whether to use TMH or not. However, anecdotal evidence suggests that some clinicians view choosing TMH over face-to-face appointments as a reluctance to fully engage.

The conclusions of this study seem rather obvious for any new development: that offering training and ongoing support would help. Training needs will differ significantly from familiar SMS messages to bespoke software, but in any case this assumes the intervention is known to be comparatively effective and desirable for all parties.

Future studies should pay specific attention to what is useful for which groups of people, including adaptations to context, before progressing to investigate implementation. Inevitably, any singular approach will leave some communities excluded.

Fundamentally, we would argue that consumers provide the ultimate litmus test of acceptability and effectiveness for any TMH modality. Co-production, from design stage to evaluation, is surely key to the success of any implementation.

## Conclusion

Using a combination of implementation strategies appeared associated with successful implementation of TMH, but it was not possible to infer conclusively causal relationships between specific types of implementation strategy and outcomes. Potentially valuable strategies to improve the implementation of TMH include providing initial training for clinicians, as well as ongoing support and consultation. Further research utilising more robust study designs to evaluate individual implementation strategies is needed to explore which specific factors can influence implementation of TMH.

## Supporting information

Appendix 1 search strategy

Appendix 2 quality appraisal

Consent form 1

Consent form 2

## Data Availability

All data produced in the present work are contained in the manuscript

## NIHR Mental Health Policy Research Unit

This paper presents independent research commissioned and funded by the National Institute for Health Research (NIHR) Policy Research Programme and conducted by the NIHR Mental Health Policy Research Unit (MHPRU). The views expressed are those of the authors and not necessarily those of NIHR, the Department of Health and Social Care or its arm’s length bodies, or other government departments.

