## Appendix 1 search strategy for "Implementation strategies for telemental health: a systematic review"

SEARCH CONDUCTED JULY 7^TH^ 20201

Pubmed 3017

WoS 5211

CINAHL 3777

EMBASE 4722

PsychINFO 4131

=20858

Duplicates removed = 14,295

PsychINFO

[*mental disorders]*

1. mental disorders/ or anxiety disorders/ or obsessive compulsive disorder/ or panic disorder/ or phobias/ or social phobia/ or bipolar disorder/ or eating disorders/ or anorexia nervosa/ or binge eating disorder/ or bulimia/ or affective disorders/ or major depression/ or exp personality disorders/ or schizophrenia/ or affective psychosis/ or catatonic schizophrenia/ or paranoid schizophrenia/ or body dysmorphic disorder/ or endogenous depression/ or reactive depression/ or recurrent depression/ or treatment resistant depression/ or atypical depression/ or self-injurious behavior/ or suicidal ideation/ or attempted suicide/
2. alzheimer's disease/ or exp dementia/
3. (affective disorder* or agoraphobi* or anorexia nervosa or anxiety or BPD or binge eat* or binging or bipolar or bulimi* or combat disorder* or compulsi* or delusion* or depersonali#ation or depressed or depression or depressive or eating disorder* or EDNOS or emotional trauma or mania or manic or mood? or neurotic or obsess* or panic or paranoi* or parasuicid* or personality disorder* or phobi* or ((post-trauma* or posttrauma*) adj stress*) or psychiatr* or psychopathol* or psychosomatic or psychotic or psychos* or PTSD or schizo* or (self adj (injur* or harm or mutilat*)) or social anxiety or suicid*).ti,id,hw.
4. Mental health/ or ((mental* or psychiatric) adj2 (health* or ill* or disorder* or diagnos?s or problem*)) .ti,ab,id.
5. mental health programs/ or crisis intervention services/ or suicide prevention centers/ or community mental health/ or community mental health centers/ or community mental health services/ or community mental health training/ or community psychiatry/ or community psychology/ or mental health services/ or preventive mental health services/ or psychiatric clinics/ or (mental health service? or CAMHS or "child and adolescent mental health service?" or psychiatry or psychology or psychotherap*).ti,ab,id.
6. (OR/1-5)

[*remote working]*

7. telecommunications/ or telepsychiatry/ or telemedicine/ or computer assisted therapy/ or telephone/ or technology/ or videoconferencing/ or internet/ or computer mediated communication/ or computers/ or exp online therapy/
8. *Answering service*.ti, id.*
9. ((mobile* or phone? or telephone? or remote* or distan* or online or virtual or electronic or email or e-mail or video or remote) adj3 (consult* or counsel* or follow up or follow-up or support* or care or interview* or monitor* or therap* or treatment? or "CBT")).ti,ab,id.
10. (telemedicine or telecare or telepsychiatry or telecommunication* or teleconference* or computer assisted therap* or teletherap* or telemental or tele-medicine or tele-care or tele-psychiatry or tele-communication* or tele-conference* or tele-therap* or tele-mental).ti,ab,id.
11. *(videoconferen* or video-conferen* or videophone? Or video-phone? or video-call* or video-based call* or video call* or video based call*).ti, ab, id.*
12. *Digital interventions/*
13. *Exp telemedicine/ or computer assisted therapy/*
14. *(OR/7-13)*

[*implementation]*

15. *(implement* or adopt* or ((research or knowledge) adj (utlili* or mobil* or transfer)) or facilitat* or barrier* or challenge*).ti,ab,id.*
16. *knowledge transfer/ or patient satisfaction/ or exp treatment barriers/ or innovation/*
17. *15 OR 16*
18. *6 AND 14 AND 17*
19. limit 18 to up=20100101-20210624

*= 5223- had a quick scan they look quite relevant.*

=4256

Wos

*=any number of characters or no other characters

$ 0 or 1 character only

? = 1 character

NEAR/10 within ten words.

“” = that exact order. Truncation ok within these.

1. **TS=("affective disorder*" or agoraphobi* or "anorexia nervosa" or anxiety or BPD or "binge eat*" or binging or bipolar or bulimi* or "combat disorder*" or compulsi* or delusion* or depersonali?ation or depressed or depression or depressive or "eating disorder*" or EDNOS or "emotional trauma" or mania or manic or mood? or neurotic or obsess* or panic or paranoi* or parasuicid* or "personality disorder*" or phobi* or ((post-trauma* or posttrauma*) NEAR/2 stress*) or psychiatr* or psychopathol* or psychosomatic or psychotic or psychos* or PTSD or schizo* or (self NEAR/1 (injur* or harm or mutilat*)) or "social anxiety" or suicid*)**
2. **TS=((mental or psychiatric) NEAR/2 (health* or ill* or disorder$ or diagnos?s or problem*))**
3. **TS=(“mental health program*” or “crisis intervention service$” or “suicide prevention” or “community mental health” or “community psychiatry” or “community psychology” or “mental health service$” or “psychiatric clinic$” or CAMHS or "child and adolescent mental health service$" or psychiatry or psychology or psychotherap* )**
4. #1 OR #2 OR #3
5. **TS=((mobile$ or phon* or telephon* or remote* or distan* or online or virtual or digital or electronic or email or e-mail or video or remote) NEAR/3 (consult* or counsel* or "follow up" or follow-up or support* or care or interview* or monitor* or therap* or treatment? or CBT))**
6. **TS=(telemedicine$ or telecare or telepsychiatry or telecommunication* or teleconference* or "computer assisted therap*" or teletherap* or telemental or tele-medicine$ or tele-care or tele-psychiatry or tele-communication* or tele-conference* or tele-therap* or tele-mental)**
7. **TS=(videoconferen* or video-conferen* or videophone$ Or video-phone$ or video-call* or "video-based call*" or "video call*" or "video based call*")**
8. #5 or #6 or #7
9. **TS=(implement* or adopt* or ((research or knowledge) NEAR/1 (utili* or mobil* or transfer)) or facilitat* or barrier$ or challeng*)**
10. #4 and #8 and #9

Limit to WOS Core collection 2010 onwards

=5211

PUB MED

**(("Dementia"[MeSH Terms] OR "Alzheimer Disease"[MeSH Terms]) OR ("Mental Health"[Title/Abstract] OR "mental problem*"[Title/Abstract] OR "mental disorder*"[Title/Abstract] OR "mental illness*"[Title/Abstract] OR "Depression"[Title/Abstract] OR "depressive disorder*"[Title/Abstract] OR "Anxiety"[Title/Abstract] OR "anxiety disorder*"[Title/Abstract] OR "phobi*"[Title/Abstract] OR "agoraphobi*"[Title/Abstract] OR "anxious"[Title/Abstract] OR "obsess*"[Title/Abstract] OR "compulsi*"[Title/Abstract] OR "panic"[Title/Abstract] OR "PTSD"[Title/Abstract] OR "post traumatic stress"[Title/Abstract] OR "posttraumatic stress"[Title/Abstract] OR "stress disorder*"[Title/Abstract] OR "psychiatr*"[Title/Abstract] OR "SMI"[Title/Abstract] OR "psycho*"[Title/Abstract] OR "schizo*"[Title/Abstract] OR "manic"[Title/Abstract] OR "mania"[Title/Abstract] OR "bipolar"[Title/Abstract] OR "personality disorder*"[Title/Abstract] OR "self-harm"[Title/Abstract] OR "self-injury"[Title/Abstract] OR "self-harm"[Title/Abstract] OR "self-injury"[Title/Abstract] OR "psychological disorder"[Title/Abstract] OR "Psychiatric illness"[Title/Abstract] OR "psychiatric disorder*"[Title/Abstract]) OR ("Anxiety Disorders"[MeSH Terms] OR "Bipolar Disorder"[MeSH Terms] OR "Feeding and Eating Disorders"[MeSH Terms] OR "Depressive Disorder"[MeSH Terms] OR "Neurotic Disorders"[MeSH Terms] OR "Personality Disorders"[MeSH Terms] OR "Psychotic Disorders"[MeSH Terms] OR "Schizophrenia"[MeSH Terms] OR "Mental Disorders"[MeSH Terms:noexp] OR "Mental Health"[MeSH Terms] OR "Mentally Ill Persons"[MeSH Terms] OR "self-injurious behavior"[MeSH Terms] OR "psychology, clinical"[MeSH Terms]) OR ("Mental Health Services"[MeSH Terms]) OR ("mental health service*"[Title/Abstract] OR "CAMHS"[Title/Abstract] OR "psychiatry"[Title/Abstract] OR "psychology"[Title/Abstract] OR "psychotherap*"[Title/Abstract]))**

**AND**

**(("Telemedicine"[MeSH Terms:noexp] OR "Remote Consultation"[MeSH Terms] OR "Distance Counseling"[MeSH Terms] OR "therapy, computer-assisted"[MeSH Terms] OR "Videoconferencing"[MeSH Terms] OR "internet-based intervention"[MeSH Terms]) OR "telemedicine"[Title/Abstract] OR "telecare"[Title/Abstract] OR "telepsychiatry"[Title/Abstract] OR "telecommunication*"[Title/Abstract] OR "teleconference*"[Title/Abstract] OR "computer assisted therap*"[Title/Abstract] OR "teletherap*"[Title/Abstract] OR "telemental"[Title/Abstract] OR "videoconferen*"[Title/Abstract] OR "video conferen*"[Title/Abstract]** OR “video-call*”[Title/Abstract] OR “Video call*”[Title/Abstract] or video-based call*[Title/Abstract] OR “video based call*”[Title/Abstract] **OR "videophone*"[Title/Abstract] OR "video phone*"[Title/Abstract]) OR (("mobile*"[Title/Abstract] OR "phone*"[Title/Abstract] OR "telephone*"[Title/Abstract] OR "remote*"[Title/Abstract] OR "distan*"[Title/Abstract] OR "online"[Title/Abstract] OR "electronic"[Title/Abstract] OR "email"[Title/Abstract] OR "e-mail"[Title/Abstract]) N2 ("consult*"[Title/Abstract] OR "counsel*"[Title/Abstract] OR "support*"[Title/Abstract] OR "interview*"[Title/Abstract] OR "monitor*"[Title/Abstract] OR "therap*"[Title/Abstract] OR "treatment*"[Title/Abstract] OR "CBT"[Title/Abstract])))]**

AND

**("implement*"[Title/Abstract] or "adopt*"[Title/Abstract] or ((research[Title/Abstract] or knowledge[Title/Abstract]) N1 (utili*[Title/Abstract] or mobil*[Title/Abstract] or transfer[Title/Abstract])) or "facilitat*"[Title/Abstract] or "barrier*"[Title/Abstract] or "challeng*"[Title/Abstract])**

Limit 2010 onwards = 3017 results

EMBASE

[*mental disorders]*

1. anxiety disorder/ or obsessive compulsive disorder/ or panic/ or phobia/ or social phobia/ or bipolar disorder/ or eating disorder/ or anorexia nervosa/ or binge eating disorder/ or bulimia/ or major affective disorder/ or minor affective disorder/ or depression/ or exp personality disorder/ or schizophrenia/ or affective psychosis/ or catatonic schizophrenia/ or paranoid schizophrenia/ or body dysmorphic disorder/ or endogenous depression/ or reactive depression/ or recurrent brief depression/ or treatment resistant depression/ or atypical depression/ or suicide/ or suicide attempt/ or suicidal ideation/ or automutilation/
2. exp dementia/
3. ("affective disorder*" or agoraphobi* or "anorexia nervosa" or anxiety or BPD or bipolar or bulimi* or compulsi* or delusion* or depersonali#ation or depressed or depression or depressive or "eating disorder*" or EDNOS or "emotional trauma" or mania or manic or neurotic or obsess* or panic or paranoi* or parasuicid* or "personality disorder*" or phobi* or ((post-trauma* or posttrauma*) adj stress*) or psychiatr* or psychopathol* or psychotic or psychos* or PTSD or schizo* or (self adj (injur* or harm or mutilat*)) or "social anxiety" or suicid*).ti,ab,kw.
4. ((mental* or psychiatric) adj2 (health* or ill* or disorder* or diagnos?s or problem*)).ti,ab,kw.
5. crisis intervention/ or community mental health/ or community mental health center/ or community mental health service/ or mental hospital/ or (mental health service or CAMHS or "child and adolescent mental health service?" or psychiatry or psychotherap*).ti,ab,kw.
6. (OR/1-5)

[*remote working]*

7. telecommunication/ or telepsychiatry/ or telemedicine/ or computer assisted therapy/ or telephone/ or videoconferencing/ or internet/
8. *Answering service*.ti, kw.*
9. ((mobile* or phone? or telephone? or remote* or distan* or online or virtual or electronic or email or e-mail or video or remote) adj2 (consult* or counsel* or follow up or follow-up or support* or care or interview* or monitor* or therap* or treatment? or "CBT")).ti,ab,kw.
10. (telemedicine or telecare or telepsychiatry or telecommunication* or teleconference* or “computer assisted therap*” or teletherap* or telemental or tele-medicine or tele-care or tele-psychiatry or tele-communication* or tele-conference* or tele-therap* or tele-mental).ti,ab,kw.
11. *(videoconferen* or video-conferen* or videophone? Or video-phone? or video-call* or “video-based call*” or “video call*” or “video based call*”).ti, ab, kw.*
12. *(OR/7-11)*

[*implementation]*

13. *(implement* or adopt* or ((research or knowledge) adj (utlili* or mobil* or transfer)) or facilitat* or barrier* or challenge?).ti,ab,kw.*
14. *Implementation research/ or patient satisfaction/*
15. *13 OR 14*
16. *6 AND 12 AND 15*
17. limit 16 to dd=20100101-20210707

*=5672*

CINAHL

| **#** | **Query** | **Limiters/Expanders** | **Last Run Via** | **Results** |
| --- | --- | --- | --- | --- |
| S14 | S5 AND S10 AND S13 | Expanders - Apply equivalent subjects  Search modes - Boolean/Phrase | Interface - EBSCOhost Research Databases  Search Screen - Advanced Search  Database - CINAHL Plus | 4,580 |
| S13 | S11 OR S12 | Expanders - Apply equivalent subjects  Search modes - Boolean/Phrase | Interface - EBSCOhost Research Databases  Search Screen - Advanced Search  Database - CINAHL Plus | 684,197 |
| S12 | (MH "Program Implementation") OR (MH "Program Evaluation") OR (MH "Patient Satisfaction") | Expanders - Apply equivalent subjects  Search modes - Boolean/Phrase | Interface - EBSCOhost Research Databases  Search Screen - Advanced Search  Database - CINAHL Plus | 122,244 |
| S11 | TI ( implement* or adopt* or ((research or knowledge) N1 (utili* or mobil* or transfer)) or facilitat* or barrier* or challenge* ) OR AB ( implement* or adopt* or ((research or knowledge) N1 (utili* or mobil* or transfer)) or facilitat* or barrier* or challenge* ) | Expanders - Apply equivalent subjects  Search modes - Boolean/Phrase | Interface - EBSCOhost Research Databases  Search Screen - Advanced Search  Database - CINAHL Plus | 598,131 |
| S10 | S6 OR S7 OR S8 OR S9 | Expanders - Apply equivalent subjects  Search modes - Boolean/Phrase | Interface - EBSCOhost Research Databases  Search Screen - Advanced Search  Database - CINAHL Plus | 129,123 |
| S9 | TI ( videoconferen* or video-conferen* or videophon* or video-phon* or video-call or video based call* or video call* or video-based call* ) OR AB ( videoconferen* or video-conferen* or videophon* or video-phon* or video-call or video based call* or video call* or video-based call* ) | Expanders - Apply equivalent subjects  Search modes - Boolean/Phrase | Interface - EBSCOhost Research Databases  Search Screen - Advanced Search  Database - CINAHL Plus | 2,438 |
| S8 | TI ( telemedicine or telecare or telepsychiatry or telecommunication* or teleconference* or “computer assisted therap*” or teletherap* or telemental or tele-medicine or tele-care or tele-psychiatry or tele-communication* or tele-conference* or tele-therap* or tele-mental ) OR AB ( telemedicine or telecare or telepsychiatry or telecommunication* or teleconference* or “computer assisted therap*” or teletherap* or telemental or tele-medicine or tele-care or tele-psychiatry or tele-communication* or tele-conference* or tele-therap* or tele-mental ) | Expanders - Apply equivalent subjects  Search modes - Boolean/Phrase | Interface - EBSCOhost Research Databases  Search Screen - Advanced Search  Database - CINAHL Plus | 8,963 |
| S7 | TI ( ((mobile* or phone* or telephone* or remote* or distan* or online or virtual or electronic or email or e-mail or video or remote) N2 (consult* or counsel* or follow up or follow-up or support* or care or interview* or monitor* or therap* or treatment* or CBT)) ) OR AB ( ((mobile* or phone* or telephone* or remote* or distan* or online or virtual or electronic or email or e-mail or video or remote) N2 (consult* or counsel* or follow up or follow-up or support* or care or interview* or monitor* or therap* or treatment* or CBT)) ) | Expanders - Apply equivalent subjects  Search modes - Boolean/Phrase | Interface - EBSCOhost Research Databases  Search Screen - Advanced Search  Database - CINAHL Plus | 36,798 |
| S6 | (MH "Telehealth") OR (MH "Telecommunications") OR (MH "Telemedicine") OR (MH "Remote Consultation") OR (MH "Telepsychiatry") OR (MH "Therapy, Computer Assisted") OR (MH "Telephone") OR (MH "Videoconferencing") OR (MH "Internet") | Expanders - Apply equivalent subjects  Search modes - Boolean/Phrase | Interface - EBSCOhost Research Databases  Search Screen - Advanced Search  Database - CINAHL Plus | 98,345 |
| S5 | S1 OR S2 OR S3 OR S4 | Expanders - Apply equivalent subjects  Search modes - Boolean/Phrase | Interface - EBSCOhost Research Databases  Search Screen - Advanced Search  Database - CINAHL Plus | 629,284 |
| S4 | TI ( mental health service* or CAMHS or "child and adolescent mental health service*" or psychiatry or psychotherap* ) OR AB ( mental health service* or CAMHS or "child and adolescent mental health service*" or psychiatry or psychotherap* ) | Expanders - Apply equivalent subjects  Search modes - Boolean/Phrase | Interface - EBSCOhost Research Databases  Search Screen - Advanced Search  Database - CINAHL Plus | 54,036 |
| S3 | (MH "Psychotherapy") OR (MH "Crisis Intervention") OR (MH "Community Mental Health Services") OR (MH "Psychiatric Service") OR (MH "Emergency Services, Psychiatric") OR (MH "Psychiatry") OR (MH "Child Psychiatry") OR (MH "Forensic Psychiatry") | Expanders - Apply equivalent subjects  Search modes - Boolean/Phrase | Interface - EBSCOhost Research Databases  Search Screen - Advanced Search  Database - CINAHL Plus | 51,198 |
| S2 | TI ( anxiety disorder* or affective disorder* or agoraphobi* or anorexia nervosa or anxiety or BPD of bipolar or bulimi* or compulsi* or delusion* or depersonalisation or depersonalization or depressed or depression or depressive or eating disorder* or EDNOS or emotional trauma or mania of manic or neurotic of obsess* or panic or paranoi* or parasuicid* or personality disorder* or phobi* or ((post-trauma* or posttrauma*) N1 stress*) or psychiatr* or psychopathol* or psychotic or psychos* or PTSD or schizo* or (self N1 (injur* or harm or mutilat*)) or social anxiety or suicid* or ((mental* or psychiatric) N2 (health* or ill* or disorder* or diagnosis or diagnoses of problem*)) ) OR AB ( anxiety disorder* or affective disorder* or agoraphobi* or anorexia nervosa or anxiety or BPD of bipolar or bulimi* or compulsi* or delusion* or depersonalisation or depersonalization or depressed or depression or depressive or eating disorder* or EDNOS or emotional trauma or mania of manic or neurotic of obsess* or panic or paranoi* or parasuicid* or personality disorder* or phobi* or ((post-trauma* or posttrauma*) N1 stress*) or psychiatr* or psychopathol* or psychotic or psychos* or PTSD or schizo* or (self N1 (injur* or harm or mutilat*)) or social anxiety or suicid* or ((mental* or psychiatric) N2 (health* or ill* or disorder* or diagnosis or diagnoses of problem*)) ) | Expanders - Apply equivalent subjects  Search modes - Boolean/Phrase | Interface - EBSCOhost Research Databases  Search Screen - Advanced Search  Database - CINAHL Plus | 483,234 |
| S1 | (MH "Anxiety Disorders+") OR (MH "Bipolar Disorder") OR (MH "Eating Disorders") OR (MH "Anorexia") OR (MH "Anorexia Nervosa") OR (MH "Bulimia") OR (MH "Bulimia Nervosa") OR (MH "Affective Disorders") OR (MH "Depression+") OR (MH "Organic Mental Disorders, Psychotic") OR (MH "Schizophrenia") OR (MH "Schizoaffective Disorder") OR (MH "Personality Disorders") OR (MH "Antisocial Personality Disorder") OR (MH "Avoidant Personality Disorder") OR (MH "Borderline Personality Disorder") OR (MH "Compulsive Personality Disorder") OR (MH "Dependent Personality Disorder") OR (MH "Narcissistic Personality Disorder") OR (MH "Passive-Aggressive Personality Disorder") OR (MH "Schizotypal Personality Disorder") OR (MH "Injuries, Self-Inflicted") OR (MH "Suicide+") OR (MH "Body Dysmorphic Disorder") OR (MH "Dementia+") OR (MH "Affective Disorders, Psychotic") OR (MH "Psychotic Disorders") OR (MH "Psychological Trauma") | Expanders - Apply equivalent subjects  Search modes - Boolean/Phrase | Interface - EBSCOhost Research Databases  Search Screen - Advanced Search  Database - CINAHL Plus | 323,242 |
