## Appendix 2 quality appraisal for "Implementation strategies for telemental health: a systematic review"

Appendix 2: Results of quality assessment

MMAT for primary research studies

| Study | Percentage of criteria met |
| --- | --- |
| Adler et al (2013) | 43% |
| Baker-Ericzén et al (2012) | 29% |
| Felker et al (2021) | 57% |
| Hensel et al (2020) | 75% |
| Lynch et al (2020) | 71% |
| Lynch et al (2021) | 82% |
| Myers et al (2020) | 100% |
| Owens et al (2016) | 57% |
| Puspitasari et al (2021a) | 71% |
| Taylor et al (2019) | 86% |

AACODS checklist for descriptive studies

|  | Authority | Accuracy | Coverage | Objectivity | Date | Significance |
| --- | --- | --- | --- | --- | --- | --- |
| Chen et al (2021) | 🗸 | 🗸 | 🗸 | 🗸 | 🗸 | 🗸 |
| Lindsay et al (2015) | 🗸 | 🗸 | 🗸 | 🗸 | 🗸 | 🗸 |
| Puspitasari et al (2021b) | 🗸 | 🗸 | 🗸 | 🗸 | 🗸 | 🗸 |
| Sharma et al (2020) | 🗸 | - | 🗸 | 🗸 | 🗸 | 🗸 |
