## Supplementary material for "Implementation strategies for telemental health: a systematic review": Consent form 1

I, Karen Machin confirm the following:

- *I am a member of the* NIHR Mental Health Policy Unit's Lived Experience Working Group
- *I consent to my name being published as part of the lived experience commentary of the research paper titled “Implementation strategies for telemental health: a systematic review”*

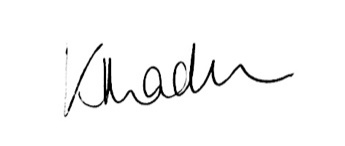
*_*

Signature for Consent

28.4.22
